# Perceptions of adolescent mothers towards adolescent repeat childbirth in Soroti District, Teso sub-region, Uganda: A phenomenological study

**DOI:** 10.1101/2022.05.07.22274735

**Authors:** Posiano Mulalu, Benon Wanume, Soita David, Dinnah Amongin, Gabriel Julius Wandawa

**Affiliations:** Department of Public Health, Faculty of Health Sciences, Mbale Campus, Busitema University, Uganda; Department of Obstetrics and Gynaecology, School of Medicine, Makerere University College of Health Sciences, Kampala, Uganda; Department of Health Policy Planning and Management, Makerere University School of Public Health, Kampala, Uganda; Department of Obstetrics and Gynecology, Faculty of Health Sciences, Busitema University, Mbale Campus, Uganda

## Abstract

The percentage of adolescent mothers aged 15 to 19 years with a repeat childbirth in Uganda (26.1%) is higher than the global estimate (18.5%). Soroti district tops Teso (a region with highest adolescent childbearing rate nationally) in adolescent childbearing. Adolescent repeat childbearing (ARC) is associated with poor health outcomes, increased risk of stillbirth, maternal and child mortality, thus a public health concern. The explanations to the high prevalence of repeat childbirth in Soroti district remain unknown.

We conducted a phenomenological study where theoretical saturation was achieved with 3 focus group discussions; each comprised of 8 respondents. Transcripts were organized and analyzed by QSR Nvivo following deductive approach.

**Results:** Adolescent marriage was viewed as a privilege, family planning methods were viewed as non-functional, man’s demand for sex was unchallengeable and non-supportive families including mistreatment of the participants by their families were perceived as risk factors for ARC.

This therefore suggests that in order to prevent repeat adolescent childbearing in Soroti district, and contribute towards the achievement of the SDG numbered three (ensure healthy lives and promote well-being for all at all ages) there is need to awaken and strengthen the implementation of the anti-teen marriage programs and policies; strengthen sexual/reproductive education including family planning programs, and addressing identified myths regarding ARC.

We also recommend further research to be done in Uganda, especially in other districts of Teso region to validate our findings.

## Introduction

Adolescent repeat childbearing (ARC) remains a global public health challenge, resulting into poor health, social and economic outcomes [1–4]. World Health Organization (WHO) defines an adolescent as a person aged 10 to 19 years of age[5]. Adolescent repeat childbearing (ARC) is defined by WHO as a second (or more) pregnancy ending in a live birth before age of 20 years [6, 7].

In 2018, it was estimated that, 44 live childbirths occurred among every 1000 girls aged 15 to 19 years globally [8]. The percentage of adolescent mothers aged 15 to 19 years of age with a repeat childbirth globally was estimated at 18.5% in 2017[9]. Worldwide, high rates of adolescent childbearing have been cited in developing countries. Approximately sixteen million girls aged 15 to 19 years give birth annually in developing countries [1, 10]. Since sub-Saharan Africa has the lowest contraceptive use globally, its adolescent childbearing rate (ACR) may rise, unless addressed [11].

There exist variations in the percentage of adolescent mothers aged 15 to 19 years of age with a repeat childbirth within the African countries, ranging from 3.7% in Rwanda to 26.1% in Uganda[9].

Nationally, 25% of Uganda’s adolescent girls have begun child bearing[12]. Higher ACR exist in rural areas of Uganda; 27% compared to the 19% in urban areas[12]. Uneducated Ugandan adolescent girls have a higher ACR estimated at 35%, compared to their counterparts who have attained Post-Primary Education, whose estimate is at 11%[12]. Adolescent childbearing is more common among adolescents from low wealth quintile, estimated at 33.5%, than adolescents from highest wealth quintile, whose childbearing rate is estimated at 15.1% [12]. In Uganda, Teso region has the highest ACR, estimated at 31.4% [12].

In Uganda, fluctuations exist in the estimation of adolescent repeat childbearing rate (ARCR). Some studies have reported 55.6% while others 26.1%[9, 13].

Childbearing among adolescent girls is associated with poor health outcomes among the adolescent girls and their children [14–16]. However, repeat adolescent childbirths are associated with more health risks for the mother and her child including increased risk of eclampsia, puerperal sepsis, systemic infections, low birth weight, preterm delivery and severe neonatal complications [6, 14, 17, 18]. Pregnancy and Adolescent childbirth complications are the leading cause of death among adolescent girls aged 15 to 19 years in developing countries [10]. Infants born as repeat birth to an adolescent girl are usually too small or too soon, increased child mortality and maternal mortality [19–23].

When an adolescent girl get more than one live birth, her potential to finish her education or even to get a job is limited [19]. Often, as adolescent girls move into new sexual relationship with the father of their second baby, there is a high likelihood of HIV/AIDs acquisition. Evidence shows that repeat pregnancies among adolescent girls are associated with high HIV prevalence compared to first time adolescent pregnancies [24]. Also, the first babies of the adolescent girls are often deprived of parental care, as the adolescent girls alter sexual partners. This negatively impacts on the child development[25]. Adolescents mothers with more than one gestation have lower self-esteem, greater susceptibility to unplanned pregnancy and increased use of drugs[26].

High ARCR coupled with the broad based nature of Uganda’s population pyramid is likely to have a negative impact on the Social, Health, and Economic indices of Uganda [27]. In the year 2014, 47.9% of Uganda’s population was aged zero to fourteen years (Uganda Bureau of Statistics, 2014b). This means that the above age group has either transformed into adolescents or is sooner transforming into adolescents. Adolescence is a very challenging stage, more especially in Uganda where half of her girls aged 15 to 19 years have ever had sex [29]; with the median age for sexual activity being 16.7 years [30]. Sometimes adolescent girls have reported men ten years older than them, as their sexual partners at the first sexual encounter [30]. Yet at-times such men have multiple sexual patterns [29]. This increases the risk of acquiring sexually transmitted infections (STI), pregnancy and its other associated outcomes among adolescent girls. The median birth Interval of twenty two months in Uganda [31] implies a high likelihood of Ugandan adolescent girls to have their repeat childbirth while still in adolescence. As a result, Uganda’s healthcare system is likely to be overwhelmed with health issues linked to ARC. Therefore, there is urgent need to prevent ARC so as to achieve the Sustainable Development Goal (SDG) number three (ensure healthy lives and promote well-being for all at all ages)[32].

Adolescent repeat childbearing (ARC) has attracted global and national attention. The Uganda government in collaboration with several non-governmental Organizations has intervened in various ways, including; development of National Adolescent policy, expansion and strengthening of youth friendly sexuality related services, enacting of laws and policies relating to adolescent sexuality, such as the defilement act, anti-teen marriage act, among others [2, 33, 34].

Despite the above interventions, a slight decline ARCR in Uganda has been seen; 58.9% in 1988/89 to 55.6% in 2016 [13]. This could be attributed to challenges faced by policy and program implementation [35]. Such challenges may include; cultural influence in the local areas [35]. Therefore, precisely, to scale down adolescent repeat childbearing rate in Soroti district, evidence based interventions are required.

## Methods and Materials

### Study area/study setting

The study was conducted in Soroti district. Soroti district has the highest percentage of adolescent mothers who have begun childbearing within Teso sub region “Table 1”.

**Table 1.**
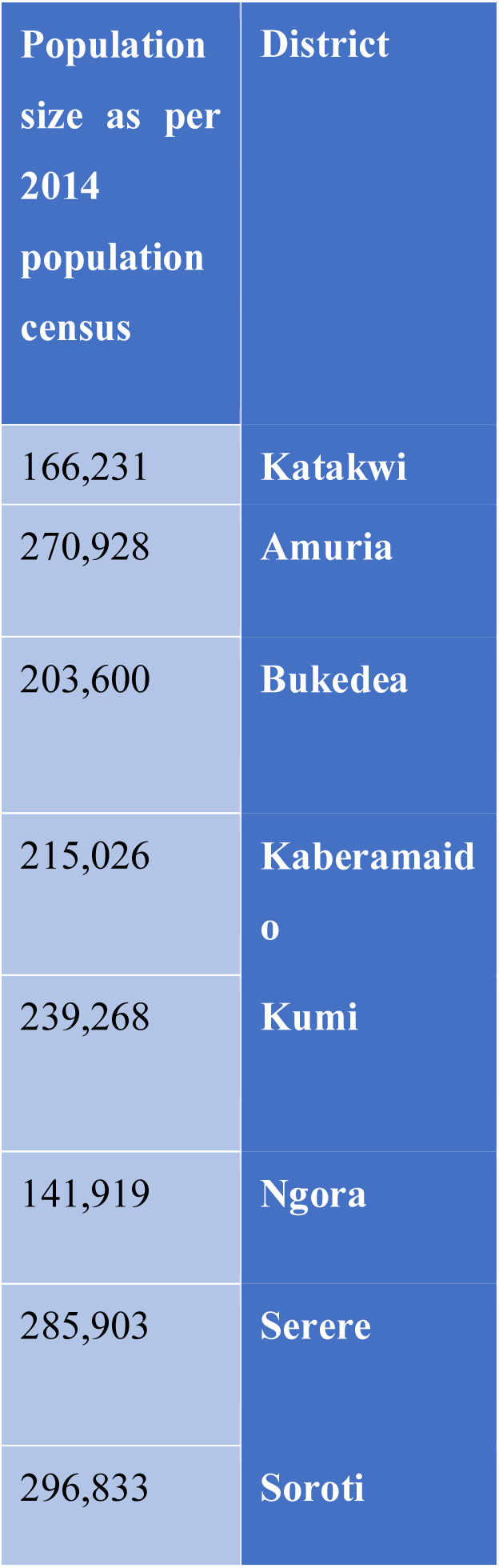
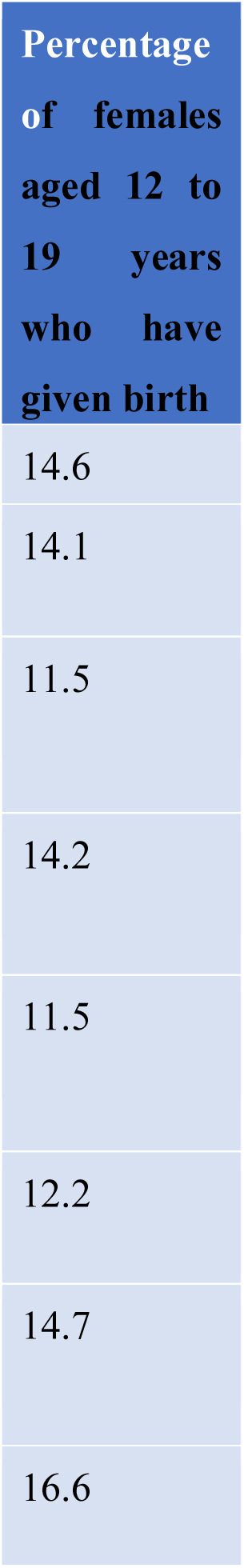
showing previous child bearing rates among adolescent females in various districts of Teso region.

The socio-cultural context in Soroti district perpetuates a high population size of people of various socio-demographics (population size of 296,833 people in 2014, of which, 13.7% are adolescent girls[28, 36]). Soroti district has an area of 2662Km. Kaberamaido, Amuria, Katakwi and Serere are its neighboring districts in the west, east, north and south respectively “Fig 1”[28]. Soroti district has three constituencies, 10 sub-counties, and 331 parishes. The major ethnic groups in Soroti are the Itesots and Kumams [28].

### Study design

This was a phenomenological study where Focus group discussions (FGDs) were conducted to collect textual data regarding the perceptions of adolescent mothers aged 15 to 19 years with repeat childbirth towards adolescent repeat childbearing.

#### Study Population

Adolescent mothers aged 15 to 19 years with repeat childbirth found in Soroti district

#### Number of interviews required to achieve theoretical saturation

Three focus group discussions were conducted among 24 adolescent mothers aged 15 to 19 years with repeat childbirth. Each focus group discussion consisted of eight participants.

#### Sampling procedure

Upon identification of the potential participant, the research team would mobilize her. The identified participant was explained to the study details, asked for consent on voluntary basis. Once she consented, she would be issued an invitation letter, indicating the date, time and venue where the interview was to take place. This was done until when all the required 8 participants were enrolled for an FGD. Prior to commencement of the FGD, consent was again sought from the participants for their participation, and to record their voices. The process was repeated to achieve the required number of participants for the next FGD. This was continued until when theoretical saturation was achieved (a point in time when no new information was generated). Theoretical saturation of the data was achieved with 3 FGDs (24 participants).

##### Inclusion Criterion

During the administration of the questionnaire, adolescent mothers aged 15 to 19 years, with a repeat childbirth, who consented to further participate in the FGDs, were included.

##### Exclusion criterion

During the administration of the questionnaire, adolescent mothers aged 15 to 19 years, with a repeat childbirth, who failed to consent to further participate in the FGDs, were excluded.

#### Data collection tool

An interview guide was used with open ended questions, and probes were asked during the interview process, depending on the way the participant answered.

#### Data Collection methods

Three focus group discussions (FDGs) were conducted. Each FDG consisted of eight participants. FDGs were conducted in a relaxed and quiet setting and in local languages. Translation back into English was done and detailed notes taken. Interviewer asked up to eight open-ended questions with follow-ups (“probes”) for clarification where it was necessary. Each focus group session lasted not less than 1½ hours.

Data collection for focus group interviews lasted for 2 days.

#### Study validity

##### Internal validity

There was translation of study into local languages (for participants who did not understand English) and re – translation back to English by research assistants.

Also, proper selection of participants with the right data, to participate ensured internal validity.

##### External validity

Findings only apply to the adolescent mothers aged 15 to 19 years with repeat childbirth found in Soroti district, Teso sub-region.

#### Data Management

##### During and after data collection

A researcher and a research assistant conducted FGDs. The researcher asked the questions in English while the research assistant translated Questions into local Languages for the participant. The research assistant also re-translated the participant’s responses from the local languages back into English for the researcher. The research assistant audio recorded, while the researcher took detailed notes. Data collected inform of audio recordings, and transcripts were safely kept in locked cardboards.

Multiple Soft copies of the data were made and stored on different hard discs and each was kept under tight privacy and confidentiality using password.

##### Data Management during data processing

Caution was taken throughout the analysis. Analyzed data was kept in the sub-folders and each was encrypted for security.

Results were merged into a master file. All files were encrypted for security purposes.

##### Data security

No personally identified information was used. Codes were given to the participants.

#### Data Analysis Plan

During the focus group discussions, detailed notes were taken. Data was analyzed for the themes. This was an iterative process beginning with data collection. N-vivo was used to manage the qualitative data which consist of transcripts. Coding of data was done. Major themes and relationship between the ideas were noted.

#### Data presentation

Results were presented inform of textual descriptions of themes generated from the qualitative data.

### Ethical consideration

Ethical clearance was sought from Mbale Regional Referral Hospital Research and Ethics Committee. Permission was thought from the DHO of Soroti district. Clearance was obtained from Busitema University, and higher degrees committee of Busitema University.

Photocopies of the approval letter from the Research and Ethics Committee and introductory letter from Dean faculty of Health sciences, Busitema University was made and each separately counter-signed by the District Health Officer Soroti district. These letters were further photocopied, and the photocopies used as introductory letters to the different leaders at various levels.

A clear and detailed description of the study protocol was given to the different stakeholders including, the leaders and study participants. An informed consent was obtained from the study participants since some who were below 18 years were emancipated minors since they had given birth.

Equally, the local council chairpersons of the villages were approached by the researcher. They were be explained to, the research details. Formal permission from the local council chairpersons was sought, through obtaining oral consent.

Data was collected from participants who consented to participate. High degree of professionalism, confidentiality and privacy was exhibited during the data collection exercise.

Participants’ names were not collected during the data collection processes, data analyses and research publication. Rather, codes were used to ensure participant confidentiality and anonymity.

Furthermore, the completed participant consent forms were kept under locked cupboards and only accessible to the study team or authorized person(s) in case need arises.

During data collection, the participants consented to have the research findings published.

### Dissemination of results

Results of the study have been availed to Busitema University, the District Health Office of Soroti, and Mbale RRH REC and orally presented to area gatekeepers/leaders from which data was collected.

### Study Limitations

Recall and Information bias were the likely limitations of the study since participants were asked about past events in their life.

External validity is a limitation to the findings of the qualitative data. Findings can only be applied to Soroti district, Teso sub-region.

### Psycho-social Support

No participant had her deep-seated emotions awakened.

### Safety of study team and Participants against COVID19

There was strict adherence to the ministry of health standard operating procedures against COVID 19. Social distancing of at-least two meters and use of hand sanitizers and masks were strictly worn by the study team.

## RESULTS, INTERPRETATION AND PRESENTATION

Theoretical saturation was attained with three focus group discussions (FGDs). Each FGD consisted of eight participants, with at least a member of the age group 15 to 19 years. FGDs were conducted in a relaxed and quiet setting and in local languages. Translation back into English was done and detailed notes taken. Interviewer asked up to eight open-ended questions with follow-ups (“probes”) for clarification where it was necessary. Social distancing and wearing of face masks were highly adhered to during the interviews, given that the study was conducted during the period of COVID-19 pandemic.

Of the 24 respondents within the 3 FGDs conducted; Seventeen respondents were married while 7 were single. Nineteen respondents ended in Primary while 5 in secondary.

Four themes emerged regarding the perceptions of adolescent mothers on adolescent repeat childbirth which predispose them to repeat childbirth. These were; participants being married, mistreatment of participants by their families, factors related to sexual intercourse, and participants having inappropriate sexual reproductive health information.

These themes are derived from the following sub-themes as shown below.

Perceived personal factors.

Throughout the three FGDs, adolescent repeat childbirth was not a surprise. They had various perceptions to explain. These included: -

Being married: Respondents viewed marriage as a privilege and a contract whose results were childbearing as some participants noted: “… I was taken to produce, so I had to produce for the man children… (Married and ended in secondary)” and another participant said; “… my husband said; that is the contract you have been brought for, so lets us produce… (Married and ended in Primary)”

Respondent’s Personal urge to have sex. Some respondents perceived sex as so sweet that it was unavoidable after the first childbirth, hence went for another attempt. A respondent noted: “…the style of having sex really prompted me to have another attempt… (Married and a primary leaver)”

Being afraid, following the first pregnancy. This partly resulted from family members’ reactions or community stigmatization. Respondents were predisposed to marriage, as they left their parent’s households for either relatives or the sexual partner’s residence. A case study is: “…I became afraid and took off with my boyfriend without my father’s knowledge… (Married and Primary leaver)” while another participant asserted that: “…some people would just whisper, “you see that girl, she is pregnant”. I would feel bad and this made me to go and marry… (Married and Primary leaver)”

Inability to reject sexual intercourse whenever the sexual partner demanded was also perceived as a likely exposure to repeat childbirth. They claim their sexual Partner’s decision is unchallengeable. An example; “…may be in need of sex, when he wants, I have to satisfy him… (a single, primary leaver)”

Initiation of sex/childbearing at an early age: Respondents viewed the practice of sexual activity/childbearing in adolescence as normal hence prompted them to have sex again, resulting into a repeat childbirth. One of the respondents said: “…I was old enough to get a baby, so I produced… (Married and Primary leaver)”

Confirmation of sexual functionality was yet another factor. Some respondents wished to find out whether the first childbirth was not an accident. Some were not sure whether they are the ones who had produced their first baby as they claimed they did not know where the first baby passed during delivery. This was mostly among respondents who never received reproductive health education. A participant noted: “…I tested sex for the first and second times to test whether I really function normally… (Single and Primary leaver)”

Loss of the first baby made some adolescent mother to have another baby. A respondent stated: “…when my first baby passed on, I had to immediately look for another … (single and ended in primary)”

Though the study findings indicate to some extent, that community and family discouraged childbirth among adolescent girls (for example; “… My clan and community members were not happy for the pregnancy, they told me that they used to tell me about education but I did not listen… (single, primary leaver)”), the respondents perceived their families, peers and communities to have had an influence on repeat childbirths.

Mistreatment of the respondents by their families came out strongly to have influenced the respondents to marry, which predisposed them to a repeat childbirth as one of the respondents noted: “…My brothers beat me up… (Married, primary leaver)”. Another case study is; “…I was denied food and slept hungry… (Married, primary leaver)”.

Non-supportive families to adolescent mother, was also seen to have influenced a repeat childbirth among the respondents. Lack of emotional or financial support or both was reported throughout the three FGDs. Adolescent Pregnancy was a burden and family shame:

“… do not bring problems for us; we do not have bastered in our family, just locomote out of this family… (Married, secondary leaver)” and another participant stated that; “…my brothers said, for us in this family we do not land for a bastered, you’re not fit to be in this family… (Married, primary leaver)”.

Adolescent pregnancy was a source of income. All the three FGDs show adolescent pregnancy as a source of dowry for the sons and money for the family members. Pregnant adolescents were rushed to the families of their sexual partners according to the findings:

“…my brother had brought a K-girl, but at home there was no-thing to pay dowry for my brother, so I went to get dowry for my brother… (Single, Primary leaver)” and another respondent said; “…clan at large was so happy, saying; if I conceive they will pay dowry so that brothers in my clan would get married… (Married, primary leaver)”

The study findings show influence of peers of adolescent mothers to have been critical. Some peers were encouraged to have a repeat childbirth: “…I was encouraged by my peers to have a repeat child; they said the children would be my supporter in old age… (Single, Primary leaver)”

The following reproductive health related factors were perceived to have influenced adolescent repeat childbirth:

According to the study findings, not using any of the family planning method was yet a strong associated with repeat childbirth. The barrier to its use was identified as lack of information about reproductive health including family planning or rejection of family planning use by the sexual partner even though the adolescent mother wanted to use. “…I did not get any information since I did not go for any antenatal visit… (Married, secondary leaver)”. Another said “…I told my husband to use family planning, but he refused to use… (Married, primary leaver)”

Failure of the family planning methods to work, even though the adolescent mother used, was equally perceived as a risk factor for adolescent repeat childbirth “…. I used family planning method, but failed to work, consequently I conceived my second baby… (Married, primary leaver)” and another respondent said; “…also, I used family planning but did not work… (Married, primary leaver)”.

Sexual partners were great influencers of adolescent repeat childbirth. Sexual partner related factors tended to overpower all the factors from the other themes. These factors included; failure of the sexual partner to use condoms, provision of adolescent mother’s need, and sexual partner’s demand for either sex or children.

“…My husband needed a second attempt of sex, so I would give him sex whenever he needed… (Married, primary leaver)”.

## DISCUSSION OF RESULTS

### Perception of adolescent mothers towards repeat childbirth

The participants perceived adolescent marriage as an opportunity. This possibly stems from the community perception of un-married adolescent mothers as failures. Some community members within Soroti district felt pity for un-married adolescent mothers. Some adolescent mothers were forced by their parents or relatives into marriages, to raise either dowry for their elder brothers or family income.

Participants claimed family planning methods failed to work as expected. This could indicate that adolescent mothers received no or inappropriate information regarding sexual and reproductive health. It implies that interventions aimed at preventing repeat adolescent childbirth need to fully provide sexual and reproductive health education to the adolescent mothers.

The various myths related to sexual satisfaction, including; participants’ personal urge to have sexual intercourse, believing that their husband’s demand for sex is unquestionable, viewing sexual intercourse at an early age as normal, and need to confirm sexual functionality must be addressed in-order to prevent repeat childbirth among adolescent mothers. This further indicates collapse of family structures such as aunties, who used to provide sexual and reproductive health information to adolescent girls.

Non-supportive families and violence towards adolescent mothers following their first childbirth was yet another perceived risk factor for adolescent repeat childbirth. This was seen to instill fear among first-time adolescent mothers resulting into elopement with their sexual partners.

No such findings have been found in Soroti district possibly because no study has ever been carried out in Soroti district to measure perception of adolescent mothers towards repeat childbirth as an end point.

Therefore, for the effectiveness of interventions aimed at preventing repeat childbirth among adolescent mothers in Soroti district, they need to be sensitive to the above perceptions.

## CONCLUSION AND RECOMMENDATION

Qualitative findings showed that adolescent mothers perceived adolescent marriages as a privilege following their first childbirth, and childbearing in adolescent marriages was to be determined by their sexual partners, and had varying myths ranging from failure of family planning methods to confirmation of sexual functionality following their first childbirth.

## Data Availability

All relevant data are within the manuscript and its Supporting Information files

## Acknowledgement

I would like to acknowledge the various stake holders of Busitema University most especially the faculty of Health Sciences; thank you for being not only academic mentors, but also parents. Special thanks go to Prof. Peter Olupot-Olupot, Mr. Okello Francis, Dr. Matovu JKB, Dr. Iramoit Jacob, Dr. Dinah Amongin, Dr. Mukone George and the entire department of Public health, Busitema University for the scholarly guidance. In a very special way, I would like to thank Dr. Wanume Benon and Dr. Soita David for the tireless academic support they extended to this study.

Gratitude goes to my wife; Joan, our children; Abigail, Ariana and Anna, for their love and support that kept me going.

## Supporting Information

Appendix

