## Appendix for "Perceptions of adolescent mothers towards adolescent repeat childbirth in Soroti District, Teso sub-region, Uganda: A phenomenological study"

### APPENDICES

#### Appendix I: Consent form:

**Consent**

**Title of the proposed study:**	Factors associated with repeat childbirth among adolescent mothers in Soroti district, Teso region, Uganda; a mixed methods study.

**Investigator:**

Mulalu Posiano (BSc. Biology), a Master of Public Health Student, Busitema University.

**Introduction:** Globally, 20% of adolescent childbirths are repeats. In Uganda, adolescent repeat childbearing rate (ARCR) is 55.6%. Adolescent childbearing rate (ACR) in Uganda is higher than global average (24.8% versus 4.4% respectively). Teso region tops Uganda, with an ACR of 31.4%. Soroti district tops Teso sub-region in adolescent childbearing. Repeat childbearing is associated with vast health, social and economic challenges. Repeat teenage births have been associated with almost threefold risk of preterm delivery and stillbirth compared to first teenage birth. The situation worsens when fathers of the first and repeat children differ; child spacing among the adolescent females is often of short interval, first born of adolescent females are often deprived of parental care since; they are often left with their grandparents. Repeat childbirth among adolescent females aged 15 to 19 years is thus a public health problem. Not only are the factors associated with repeat childbirth among adolescent girls in Soroti district unknown, but also is the proportion of adolescent mothers with repeat childbirth. Hence, interventions aimed at reducing repeat childbirth among adolescent mothers have not been informed by empirical data.

**Objective:** To determine the predictors of repeat childbirths among adolescent girls aged 15 to 19 in Soroti district, Teso sub-region.

**Method:** A cross-sectional study involving mixed methods of data collection will be conducted among 422 adolescent mothers aged 15 t0 19 years in Soroti district, Teso sub-region, Uganda. Quantitatively, interviewer-administered structured interviews will be conducted with 422 adolescent mothers in Soroti district. Data will be collected on the demographics of adolescent mothers and their sexual partners, socio-economics of the adolescent mothers and their sexual partners, factors related to the family of adolescent mothers and factors related to the peers of adolescent mothers. Quantitative data shall be analyzed at Univariate, Bivariate and Multivariate level using logistic regression. For the qualitative component, 4 focus group discussions (FGDs) will be conducted among 32 adolescent mothers with a repeat childbirth or pregnancy. The transcripts and audio recordings will be organized and prepared for analysis using QSR Nvivo following the deductive approach.

**Potential Contribution:** The study will generate knowledge, and inform policy makers and interventions about the factors associated with repeat childbirth among adolescent mothers aged 15 to 19 years in Soroti district. The study will aid in planning and budgeting to prevent repeat childbirth among adolescents mothers.

**Who will participate in the study?**

In the focus group discussion, the first thirty-two adolescent mothers with a repeat childbirth or pregnancy in Soroti district who consent to participate in the study will be included.

**Risks/Discomforts:**

There is no potential physical risk in this study except a recap of the bad emotional experiences which may arise as you remember the history of bad moments that you might have experienced. In case, such emotions arise, you will be counselled by the researcher since he is a trained teacher with professional training in counselling.

**Confidentiality:**

High confidentiality level will be adhered to throughout every stage of this research. Codes shall be used instead of your names. The codes shall tally with the codes in the registers at the point of obtaining health services. Only the ethical bodies and Busitema University may have access to any personally sensitive information concerning you if required.

**Alternatives:**

Participation in this study is optional. Even if you accept to take part in the study, you can exit the study at any time without informing any research member team.

**Compensation for participation in the study:**

Since the research does not involve any risk, there is no anticipated injury hence no compensation to any participant shall be given.

**Reimbursement:** Participants in the FGDs will receive refreshment during the participation and a transport refund of two thousand shillings only. However, during the administration of the questionnaire, nothing will be given to the research participant, as the research is sponsored by the student.

**Questions:**

If case of any question regarding this research, you can reach out to the investigator by; calling on +256-759-996-288 or emailing the investigator at;

**Questions about participants rights:**

In case of any inquiry regarding your rights as a participant, you can reach out to the investigator by; calling on +256759996288 / 0778381350 or emailing the investigator at;, or the supervisor Dr. Wanume Benon on cellphone +256-77-479249.

**Statement of voluntariness:**

Participation in this study is voluntary and you may join on your own free will. You have a right to withdraw from the study at any time without penalty.

**Statement of consent:**

If you have fully understood the above, then fill the STATEMENT OF CONSENT below;

**STATEMENT OF CONSENT/ASSENT**

........................................................................... has described to me what is going to be done, the risks, the benefits involved and my rights regarding this study. I understand that my decision to participate in this study will not alter my usual medical care. In the use of this information, my identity will be concealed. I am aware that I may withdraw at any time. I understand that by signing this form, I do not waive any of my legal rights but merely indicate that I have been informed about the research study in which I am voluntarily agreeing to participate and have accept the study finding published. A copy of this form will be provided to me.

Name of Participant ……………………Signature ………………Date …………………

Name of Interviewer……… ……………………Signature ………………Date …………………

#### Appendix II: Interview guide

0. What sexual reproductive health services have you ever received when you were;
(a) Pregnant for your first baby
(b) After delivery of your first baby
1. How have you been doing financially after you had your first baby (Hint: Focus is on ; who finances, whether financial difficulty were experienced, how they were dealt with)
2. How did each of the following (if applicable) category of people treat you following your first childbirth?
(a) father
(b) mother
(c) brothers
(d) sisters
(e) community
3. How did you experience your first delivery?

(Hint; Focus on where they delivered, any challenge they experienced)

4. Did your peers encourage you to have a repeat childbirth? If yes, how did?
5. Was your sexual partner influential in having a repeat childbirth? If yes, how?
6. Could there be cultural factors that encouraged you to have another baby? If yes, what are they?
7. Which other factors made you to have another baby?

Appendix III: Map of Uganda showing study area

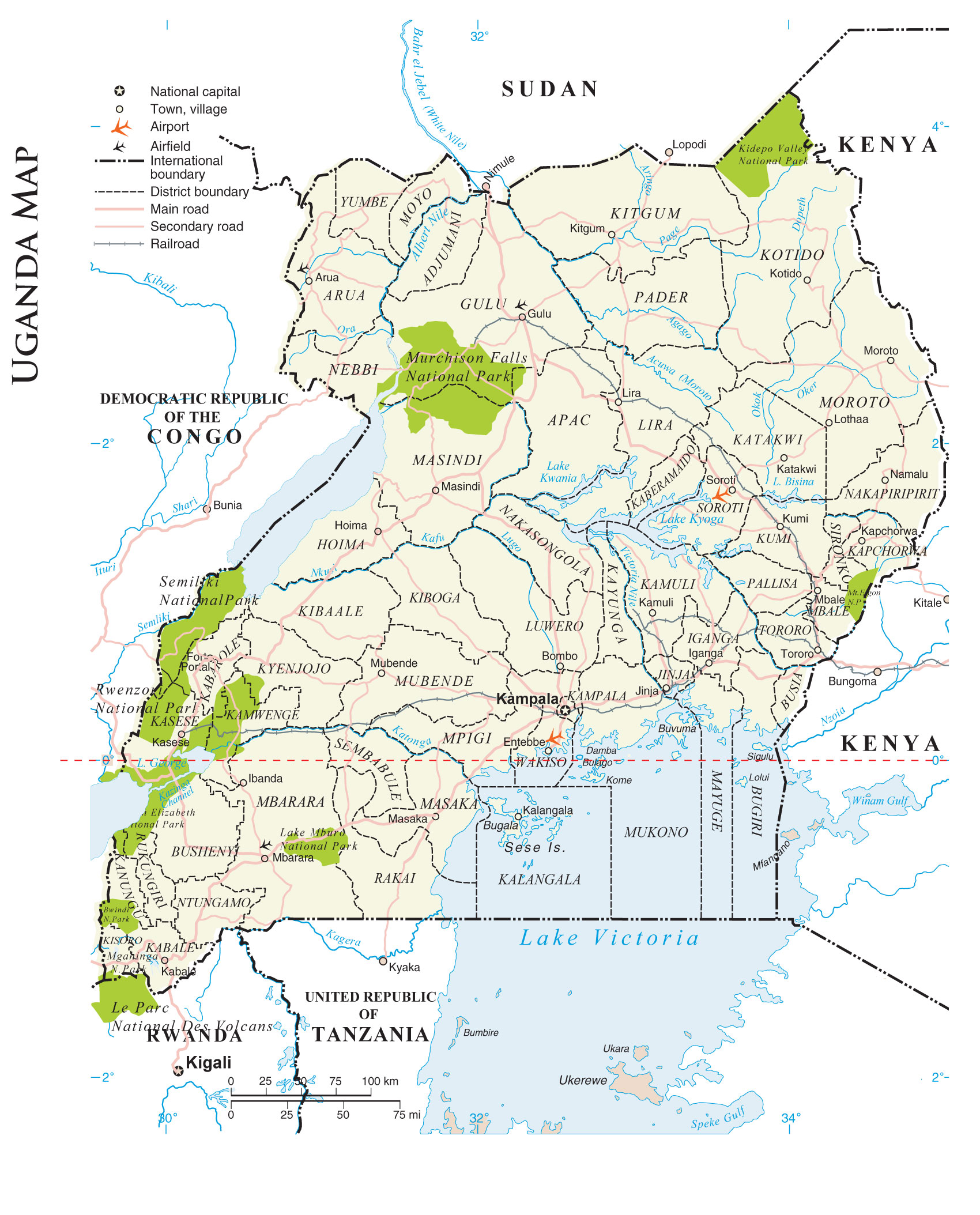

#### Appendix IV: Code book for qualitative findings

Perceptions of Adolescent mothers towards repeat childbirth in Soroti district, Uganda

Codes

| Name | Description | Files | References |
| --- | --- | --- | --- |
| 1: Personal factors related to adolescent repeat childbirth | According to the adolescent mothers, the view the following individual related factors as what could have made them to have another baby before they could jump out of adolescence | 3 | 117 |
| Being married | Marriage is seen as a contract whose results are children, according to the adolescent mothers. | 3 | 35 |
| Respondent had low income | Some respondents did not have anything that generates income; others could rely on entirely their sexual partners while others could subsidize their low earnings with what the partner could provide. | 3 | 25 |
| Respondent’s Personal urge to have sex again /another baby | Following the birth of the first baby, the adolescent mother wanted to have either sex or another baby or both due to various reasons such as style of playing sex was interesting among others | 3 | 22 |
| Fear | Adolescent mother feared to be at home following her first pregnancy either due to the community reaction or family reaction or both when they learnt of her first pregnancy | 3 | 11 |
| Respondent felt better after her first childbirth | The adolescent mother had no major challenge after the first childbirth, which encouraged her to have sex again | 2 | 9 |
| Inability to reject sexual intercourse whenever the sexual partner demanded | The mother could not resist sex when the sexual partner demanded. With or without her consent, she had to serve the sexual partner. This was reported mostly from married respondents and respondents who ended in Primary | 3 | 7 |
| Perceived early age for sex initiation | The adolescent mother had feeling the sex must be started when she is below 20 years | 3 | 5 |
| Confirmation of functionality | The adolescent mother wished to confirm whether she functions well, perhaps the first child came accidently. | 1 | 1 |
| Early initiation of coitus after first childbirth | Adolescent mother had sex soon after delivery of the first baby | 1 | 1 |
| Loss of first baby | The adolescent mother lost the first baby, thus had to get another baby | 1 | 1 |
| 2. Perceived family, peer and community factors related to adolescent repeat childbirth | The way the community, family members and peers reacted towards the first pregnancy of the respondents, and after her first childbirth was perceived by the respondent to have encouraged her to marry or go with other men hence had a repeat childbirth. | 3 | 136 |
| Non supportive family, for example no emotional support, no financial support | Family members did not support the adolescent mother when she became pregnant | 3 | 50 |
| Peers' encouragement | Peers of the adolescent mother encouraged her to have another baby | 3 | 24 |
| Being physically insulted/mistreated | The adolescent mother was mistreated following her first pregnancy, being labelled as a bastered, useless among other descriptions | 3 | 21 |
| Community stigmatization | Community was perceived as being a stigmatizing community, which scared the adolescent mother to stay in such a community. The adolescent mother could be labelled as a failure, among others | 3 | 21 |
| Adolescent childbearing as income source | Parents, clan members and relatives saw a pregnant adolescent mother as a source of income. She was taken for marriage in exchange for dowry, hence boys in a home were assured of getting married | 3 | 10 |
| Community discourages adolescent childbearing | Generally, the community discourages childbearing among adolescents | 3 | 10 |
| 3: Perceived reproductive health factors related to adolescent repeat childbirth | The following reproductive health related factors were seen to have been associated with adolescent repeat childbirth | 3 | 36 |
| No family planning | The adolescent mother did not use family planning methods after her first childbirth | 3 | 14 |
| No reproductive health education received | Adolescent mothers did not receive sex education | 3 | 13 |
| Failure of family planning methods | The adolescent mother used any family planning method but the family planning method(s) did not work | 1 | 5 |
| Never went for abortion | Adolescent mother did not abort the pregnancy despite being encouraged to abort | 1 | 2 |
| Never went for antenatal visits | Adolescent mothers did not go for antenatal visits | 2 | 2 |
| 4. Perceived sexual partner factors related to adolescent repeat childbirth | According to the adolescent mother, how her sexual partner perceives repeat childbirth | 3 | 51 |
| Need for a baby | The sexual partner wanted a baby | 3 | 24 |
| Providing necessities to adolescent mother | The sexual partner continued to support the adolescent mother. He would provide for her needs. | 3 | 14 |
| Demand for sex | The sexual partner demanded for sex from the adolescent mother, which she had to give, claiming she had been taken for that cause, or the man had given her some sort of material possession in exchange for sex | 3 | 8 |
| Refusal to use a condom during sexual intercourse | Sexual partner did not want to use a condom and the respondent had no objection over that. | 1 | 3 |
| Being a good man | The adolescent mother saw her sexual partner as a good man, hence rewarded him with another baby | 2 | 2 |

#### Appendix V: Coding matrix Query

| Themes | Socio-demographics of respondents | | | | | | | | | | |
| --- | --- | --- | --- | --- | --- | --- | --- | --- | --- | --- | --- |
|  | Age in years | | | | | Marital status | | Education level | | Tribe | |
|  | 15 | 16 | 17 | 18 | 19 | Single | Married | Primary | Secondary | Itesot | Kumam |
| 1: Personal factors related to adolescent repeat childbirth | 11 | 10 | 23 | 49 | 19 | 35 | 77 | 95 | 17 | 37 | 75 |
| - Personal wish to confirm whether she functions | 0 | 0 | 0 | 1 | 0 | 1 | 0 | 1 | 0 | 0 | 1 |
| - Early initiation of coitus after first childbirth | 0 | 1 | 0 | 0 | 0 | 0 | 1 | 1 | 0 | 0 | 1 |
| - Inability to refuse sexual intercourse when a man wants | 1 | 0 | 2 | 2 | 2 | 3 | 4 | 7 | 0 | 2 | 5 |
| - Development of fear to remain at home following the first pregnancy | 0 | 1 | 2 | 6 | 2 | 3 | 8 | 10 | 1 | 3 | 8 |
| - Felt better after my first childbirth | 2 | 1 | 2 | 4 | 0 | 4 | 5 | 8 | 1 | 2 | 7 |
| - Loss of first baby | 1 | 0 | 0 | 0 | 0 | 1 | 0 | 1 | 0 | 0 | 1 |
| - I had low income | 3 | 2 | 5 | 11 | 4 | 8 | 17 | 19 | 6 | 8 | 17 |
| - Being married | 2 | 3 | 7 | 16 | 7 | 8 | 27 | 32 | 3 | 12 | 23 |
| - It was right age to have children | 1 | 0 | 1 | 1 | 2 | 3 | 2 | 3 | 2 | 2 | 3 |
| - Personal urge to have sex following first childbirth | 1 | 2 | 5 | 10 | 4 | 5 | 17 | 17 | 5 | 10 | 12 |
| 2. Perceived family, peer and community factors related to adolescent repeat childbirth | 9 | 14 | 24 | 46 | 19 | 34 | 78 | 87 | 24 | 48 | 63 |
| - Adolescent childbearing is seen as a source of income | 0 | 2 | 5 | 3 | 0 | 3 | 7 | 9 | 1 | 6 | 4 |
| - Community discourages adolescent childbearing | 0 | 1 | 1 | 7 | 1 | 2 | 8 | 7 | 3 | 4 | 6 |
| - Adolescent being mistreated when she conceived at home | 3 | 2 | 4 | 9 | 4 | 7 | 15 | 14 | 7 | 10 | 11 |
| - Non supportive family; such as no emotional and financial support when she conceived | 5 | 5 | 11 | 21 | 9 | 13 | 38 | 39 | 11 | 22 | 28 |
| - Non-supportive communities, for example talking about the pregnancy of the adolescent mother | 2 | 3 | 5 | 8 | 3 | 8 | 13 | 16 | 5 | 8 | 13 |
| - Peers' encouragement to have another baby following her first childbirth | 1 | 3 | 3 | 12 | 5 | 9 | 15 | 17 | 7 | 10 | 14 |
| 3: Perceived reproductive health factors related to adolescent repeat childbirth | 3 | 3 | 6 | 14 | 5 | 10 | 21 | 27 | 4 | 9 | 22 |
| - Failure of the planning methods to work | 1 | 1 | 2 | 0 | 1 | 3 | 2 | 5 | 0 | 0 | 5 |
| - Never carried out abortion | 1 | 0 | 1 | 0 | 0 | 2 | 0 | 2 | 0 | 1 | 1 |
| - Never went for antenatal visits when she was pregnant for the first baby | 0 | 0 | 1 | 0 | 1 | 0 | 2 | 1 | 1 | 2 | 0 |
| - Never knew the different family planning methods | 0 | 0 | 1 | 9 | 4 | 2 | 12 | 10 | 4 | 5 | 9 |
| - Did not receive reproductive health education | 1 | 2 | 2 | 8 | 0 | 4 | 9 | 11 | 2 | 3 | 10 |
| 4. Perceived sexual partner factors related to adolescent repeat childbirth | 3 | 7 | 9 | 23 | 6 | 13 | 35 | 38 | 10 | 16 | 32 |
| - Being a good man | 0 | 0 | 1 | 1 | 0 | 0 | 2 | 2 | 0 | 1 | 1 |
| - Demanded for sex | 1 | 1 | 2 | 2 | 2 | 3 | 5 | 8 | 0 | 2 | 6 |
| - Needed a baby | 1 | 4 | 6 | 10 | 3 | 5 | 19 | 16 | 8 | 13 | 11 |
| - Provided necessities to adolescent mother | 1 | 3 | 0 | 8 | 2 | 4 | 10 | 12 | 2 | 1 | 13 |
| - He refused to use condom when having sex | 0 | 0 | 0 | 3 | 0 | 1 | 2 | 3 | 0 | 0 | 3 |
